# Benefit-risk analysis of health benefits of routine childhood immunisation against the excess risk of SARS-CoV-2 infections during the COVID-19 pandemic in Africa

**DOI:** 10.1101/2020.05.19.20106278

**Authors:** Kaja Abbas, Simon R Procter, Kevin van Zandvoort, Andrew Clark, Sebastian Funk, LSHTM CMMID CMMID COVID-19 Working Group, Tewodaj Mengistu, Dan Hogan, Emily Dansereau, Mark Jit, Stefan Flasche

## Abstract

**Background:** National immunisation programmes globally are at risk of suspension due to the severe health system constraints and physical distancing measures in place to mitigate the ongoing COVID-19 pandemic. Our aim is to compare the health benefits of sustaining routine childhood immunisation in Africa against the risk of acquiring SARS-CoV-2 infections through visiting routine vaccination service delivery points.

**Methods:** We used two scenarios to approximate the child deaths that may be caused by immunisation coverage reductions during COVID-19 outbreaks. First, we used previously reported country-specific child mortality impact estimates of childhood immunisation for diphtheria, tetanus, pertussis, hepatitis B, *Haemophilus influenzae* type b, pneumococcal, rotavirus, measles, meningitis A, rubella, and yellow fever (DTP3, HepB3, Hib3, PCV3, RotaC, MCV1, MCV2, MenA, RCV, YFV) to approximate the future deaths averted before completing five years of age by routine childhood vaccination during a 6-month COVID-19 risk period without catch-up campaigns. Second, we analysed an alternative scenario that approximates the health benefits of sustaining routine childhood immunisation to only the child deaths averted from measles outbreaks during the COVID-19 risk period. The excess number of infections due to additional SARS-CoV-2 exposure during immunisation visits assumes that contact reducing interventions flatten the outbreak curve during the COVID-19 risk period, that 60% of the population will have been infected by the end of that period, that children can be infected by either vaccinators or during transport and that upon child infection the whole household would be infected. Country specific household age structure estimates and age dependent infection fatality rates are then applied to calculate the number of deaths attributable to the vaccination clinic visits. We present benefit-risk ratios for routine childhood immunisation alongside 95% uncertainty range estimates from probabilistic sensitivity analysis.

**Findings:** For every one excess COVID-19 death attributable to SARS-CoV-2 infections acquired during routine vaccination clinic visits, there could be 84 (14-267) deaths in children prevented by sustaining routine childhood immunisation in Africa. The benefit-risk ratio for the vaccinated children, siblings, parents or adult care-givers, and older adults in the households of vaccinated children are 85,000 (4,900 - 546,000), 75,000 (4,400 - 483,000), 769 (148 - 2,700), and 96 (14 - 307) respectively. In the alternative scenario that approximates the health benefits to only the child deaths averted from measles outbreaks, the benefit-risk ratio to the households of vaccinated children is 3 (0 - 10) under these highly conservative assumptions and if the risk to only the vaccinated children is considered, the benefit-risk ratio is 3,000 (182 - 21,000).

**Interpretation:** Our analysis suggests that the health benefits of deaths prevented by sustaining routine childhood immunisation in Africa far outweighs the excess risk of COVID-19 deaths associated with vaccination clinic visits, especially for the vaccinated children. However, there are other factors that must be considered for strategic decision making to sustain routine childhood immunisation in African countries during the COVID-19 pandemic. These include logistical constraints of vaccine supply chain problems caused by the COVID-19 pandemic, reallocation of immunisation providers to other prioritised health services, healthcare staff shortages caused by SARS-CoV-2 infections among the staff, decreased demand for vaccination arising from community reluctance to visit vaccination clinics for fear of contracting SARS-CoV-2 infections, and infection risk to healthcare staff providing immunisation services as well as to their households and onward SARS-CoV-2 transmission into the wider community.

**Funding:** Gavi, the Vaccine Alliance and Bill & Melinda Gates Foundation (OPP1157270)

**Research in context:** *Evidence before the study:* National immunisation programmes globally are at risk of disruption due to the severe health system constraints caused by the ongoing COVID-19 pandemic and the physical distancing measures to mitigate the outbreak. The decrease in vaccination coverage increases the proportion of susceptible children at risk of increased morbidity and mortality from vaccine-preventable disease outbreaks. Outbreaks of vaccine preventable disease have been observed during previous interruptions to routine immunisation services during an ongoing infectious disease epidemic, such as during the 2013-2016 Ebola outbreak in West Africa, when most health resources were shifted towards the Ebola response which led to decreasing vaccination coverage and consequently outbreaks of measles and other vaccine-preventable diseases.

*Added value of this study:* We estimated the benefit-risk ratio by comparing the deaths prevented by sustaining routine childhood immunisation for diphtheria, tetanus, pertussis, hepatitis B, *Haemophilus influenzae* type b, pneumococcal, rotavirus, measles, meningitis A, rubella, and yellow fever vaccines with the excess COVID-19 deaths associated with vaccination clinic visits. The benefit of routine childhood immunization programmes in all the 54 countries of Africa is higher than the COVID-19 risk associated with these vaccination clinic visits.

*Implications of all the available evidence:* Routine childhood immunisation programmes should be safeguarded for continued service delivery and prioritised for the prevention of infectious diseases, as logistically possible, as part of delivering essential health services during the COVID-19 pandemic in Africa. The current immunisation service models will require adaptation, including physical distancing measures, personal protective equipment, and good hygiene practices for infection control at the vaccination clinics, and have to be complemented by new immunisation service models for sustaining routine childhood immunisation in the African countries during the COVID-19 risk period.

## Introduction

Vaccines have substantially improved health and reduced mortality, particularly among children in low-income countries.^1–3^ Access to vaccines in these countries accelerated after the formation of Gavi, the Vaccine Alliance in 2000.^4^ This access needs to be sustained to further advance the public health gains and maintain progress towards goals such as the elimination of polio, measles, rubella, and maternal tetanus.^5^ The World Health Organization has launched its Immunization Agenda 2030 strategy in order to accelerate progress towards equitable access and use of vaccines over the new decade.^6^ However, ensuring everyone has access to immunisation services has proved challenging, with a quarter of children in the Africa region not receiving three doses of diphtheria-tetanus-pertussis (DTP3) in 2018.^7^ This is now further challenged by the coronavirus disease 2019 (COVID-19) pandemic,^8^ which has necessitated physical distancing measures to mitigate or delay the coronavirus epidemic that threatens to overwhelm health care systems^9,10^

The severe acute respiratory syndrome coronavirus 2 (SARS-CoV-2) emerged in December 2019 causing cases of COVID-19 in Wuhan, China.^11^ As of June 23, 2020, there were 8,993,659 confirmed cases and 469,587 confirmed deaths affecting 216 countries and territories.^12^ All African countries have reported cases with the majority reporting local transmission and rapidly rising case numbers.^13^ The prevention and control measures to suppress and mitigate the COVID-19 outbreak in Africa during the upcoming months will place immense pressures on the national health systems in their provision of essential health services, including the Expanded Programme on Immunization (EPI) and routine vaccination of infants.^14^

On March 26, 2020, the World Health Organization and the Pan American Health Organization issued guidance on the operation of immunisation programmes during the COVID-19 pandemic.^15,16^ The guidance advises for temporary suspension of mass vaccination campaigns and a risk-benefit assessment to decide on conducting outbreak response mass vaccination campaigns, while routine immunisation programmes should be sustained in places where essential health services have operational capacity of adequate human resources and vaccine supply while maintaining physical distancing and other infection control measures.

Our aim is to compare the health benefits of sustaining routine childhood immunisation in Africa against the risk of acquiring SARS-CoV-2 infections through visiting routine vaccination service delivery points. Specifically, we conducted a benefit-risk analysis of vaccine-preventable deaths averted by sustaining routine childhood immunisation in comparison to excess COVID-19 deaths from SARS-CoV-2 infections acquired by visiting routine vaccination service delivery points.

## Methods

### Assumptions

We assess the benefit and risk of continued routine childhood immunisation during the COVID-19 pandemic in all 54 African countries. We focus on the delivery of infant immunisation at: (i) 6, 10 and 14 weeks of age for diphtheria, tetanus and pertussis (DTP), polio, hepatitis B (HepB), *Haemophilus influenzae* type b (Hib), *Streptococcus pneumoniae*, rotavirus (hereafter called EPI-1); (ii) 9 months for measles (MCV1), rubella (RCV1), *Neisseria meningitidis* serogroup A (MenA), yellow fever (YFV) (hereafter EPI-2); and (iii) 15-18 months for the second dose of measles (MCV2; EPI-3). The target age for MenA routine immunization varies by country and is given along with the first or second dose of measles – 9 months in Central African Republic, Chad, Côte d’Ivoire, Mali, Niger, and Sudan; 15 months in Burkina Faso; and 18 months in Ghana.^17^ We did not consider Bacillus Calmette-Guérin (BCG) or HepB birth dose because they are recommended for administration shortly after birth and thus were assumed not to require an additional vaccination visit, albeit home births or delayed administration may be common in some parts of Africa.

During the period of SARS-CoV-2 circulation, we assume that contact-reducing measures are in place and that while those measures fail to contain the outbreak, they will be able to substantially flatten the epidemic curve. However with gradual easing of interventions and in the absence of a vaccine, SARS-CoV-2 transmission will infect around 60% of the population. In both other qualitatively different scenarios (uncontrolled epidemic or successful containment) sustaining vaccination as far as possible would be the largely obvious choice as doing so would not substantially affect the risk of SARS-CoV-2 infection.

We assume that the risk from COVID-19, and hence the potential disruption to the health services including routine childhood vaccination lasts for 6 months. The main analyses consider the impact of continuation of all five immunisation clinic visits in comparison with the risk for COVID-19 disease in the vaccinees household as a result of attending the vaccine clinic, tracking the health benefits from immunisation among the vaccinated children until five years of age.

### Benefits of sustained routine childhood immunisation

We used the health impact estimates provided by Li et al for vaccines against hepatitis B, *Haemophilus influenzae* type b, measles, *Neisseria meningitidis* serogroup A, *Streptococcus pneumoniae*, rotavirus, rubella, and yellow fever.^3^ For the health impact of vaccines against diphtheria, tetanus and pertussis (DTP), we calculated basic estimates for the annual number of deaths averted per 1000 vaccinated children by DTP in Africa based on global annual DTP3 vaccine impact estimates from 1980 to 2013.^18^ We did not include polio vaccine preventable mortality into our estimates. Antigen-specific estimates of per-capita deaths averted by vaccination were unavailable for 9 countries, and were approximated to the mean estimates of other countries with available data. Country and antigen-specific levels of routine vaccination coverage are assumed to be the same level as 2018 for 2020.

The child deaths averted by routine vaccination during a 6-month suspension period of immunisation are the product of country and antigen-specific estimates of per-capita deaths averted by vaccination from the time of vaccination until 5 years of age,^3,18^ country-specific population estimates of the vaccinated cohort,^19^ country and antigen-specific official country reported estimates of vaccination coverage,^20^ and the suspension period of immunisation.

We considered two scenarios – high-impact and low-impact, for approximating the impact of sustaining routine childhood immunisation during the COVID-19 pandemic. In the high-impact scenario, we approximate the impact of sustaining routine childhood immunisation with the estimates of vaccination impact for a 6-month cohort in 2020. Hence, this scenario assumes that the suspension of immunisation will result in a cohort of unvaccinated children who have the same risk of disease as children in a completely unvaccinated population, and their vulnerability persists until they are 5 years old, i.e. no catch-up campaign will be conducted at the end of the SARS-CoV-2 outbreak. Because of herd protection and likely catch-up activities at the end of a potential disruption of immunisation services, this high-impact scenario very likely overestimates the negative impact of suspending immunisation services for a short period of time.

In contrast, the low-impact scenario attempts to estimate a lower bound on the expected number of deaths due to disruptions to routine childhood immunisation services. We assume that in the absence of immunisation, herd immunity would protect children missing out on vaccination from all diseases with the exception of measles, and that vaccination through catch-up campaigns would close measles immunity gaps immediately following the 6 month COVID-19 disruption period. This scenario is implemented as illustrated by the following example. In a country with 80% routine measles vaccine coverage, the inter-epidemic period of measles outbreaks is about 4 years.^21^ The suspension of the routine vaccination programme for 6 months would correspond to an accumulation of susceptibles equivalent to 30 months in normal times, thus shrinking the inter-epidemic period to 2 years. In the absence of supplementary immunisation activities this would yield a 25% chance that an outbreak starts during the 6 months of suspension. Further, the physical distancing interventions in place to mitigate the COVID-19 risk may decrease that outbreak probability by an additional 50%. Thereby, there is a 12.5% (25% * 50%) chance of a measles outbreak during the 6-month suspension period. In this low-impact scenario, the health impact of routine childhood immunisation only includes a 12.5% proportion of the health benefits derived from measles vaccination while excluding the health impact of the other vaccines.^22^

Supplementary immunisation activities for 15 African countries in 2020 are either currently postponed or of unknown status and reflect higher risk for measles outbreaks in comparison to the low-impact scenario used in this study.^23^ If routine childhood immunisation programmes are also suspended, the further decline in vaccination coverage enhances the risk of measles outbreaks in the near future.

### Excess risk of COVID-19 disease from sustained routine childhood immunisation

We assume that in the coming months that African countries will experience SARS-CoV-2 spread similar to that observed in non-African countries affected earlier in the pandemic which were unable to contain the virus. Particularly, we assume that the warmer climates in Africa will not notably reduce the transmissibility of SARS-CoV-2.^24,25^

The risk of COVID-19 depends on exposure probability to SARS-CoV-2 and progression to disease. For this analysis, we only consider the infection fatality risk for COVID-19 and ignore other potentially severe health outcomes. We model the additional SARS-CoV-2 exposure risk for the vaccinated child, their carer, and household members as a result of contact with the vaccinator and other community members during travel to the vaccine clinic. The simulation parameters for SARS-CoV-2 infection dynamics are shown in Appendix A1 based on the Reed-Frost epidemic model,^26^ and the COVID-19 risk model is described in more detail in Appendix A2. We use the country-specific household age composition to approximate the age distribution in households at risk of SARS-CoV-2 infection given that one of the household members is a child who has been vaccinated, and is further elaborated in Appendix A3.^27^ We apply age-stratified infection fatality risk for SARS-CoV-2 using estimates obtained from reported cases and their severity in China in combination with the proportion of asymptomatic infections estimated among international residents repatriated from China.^28^ For children, we used the reported risks for ages 0-9 years, for adults the risk for ages 30-39 years, and for older adults over 60 the risk for ages 60 years and above (see Table A3).

### Sensitivity analyses

We conducted a probabilistic sensitivity analysis by conducting 4000 simulation runs to account for the uncertainty around the parameters governing the SARS-CoV-2 infection model, as well as the reported uncertainty ranges for the infection fatality rate estimates (modelled using a gamma distribution), and the vaccine preventable mortality estimates (modelled using a lognormal distribution), and assessed their impact on our findings.

The program code and data for the benefit-risk analysis conducted in this study is accessible on GitHub (https://github.com/vaccine-impact/epi_covid). All analyses were done using R 3.6.0.^29^ All data were from secondary sources in the public domain, and ethics approval was thereby not required.

## Results

In the high-impact scenario, we estimate that the current routine childhood immunisation programme (DTP, HepB, Hib, PCV, RotaC, MCV, RCV, MenA, YFV) in Africa during a 6 months period in 2020 would prevent 702,000 (635,000 - 782,000) deaths in children from the time of vaccination until they are 5 years old. About one third of averted deaths are attributable to measles and another third to pertussis. Immunisation during the three EPI-1 visits for DTP3, HepB3, Hib3, PCV3, and RotaC will prevent 471,000 (406,000 - 548,000) deaths, immunisation during EPI-2 visit for MCV1, RCV1, MenA, and YFV will prevent 220,000 (205,000-236,000) deaths, and immunisation during EPI-3 for MCV2 will prevent 10,300 (9,400 - 11,200) deaths among children until they are 5 years old (see Table 1). One-third of the deaths prevented by routine childhood vaccination are in Nigeria, Ethiopia, Democratic Republic of Congo, and Tanzania (see Table 2).

**Table 1:** Vaccine antigen specific benefits and risks of sustaining routine childhood immunisation in Africa. The benefit-risk ratio estimates (median estimates and 95% uncertainty intervals) show the child deaths averted by sustaining routine childhood immunisation in Africa per COVID-19 death attributable to excess SARS-CoV-2 infections acquired through visiting routine vaccination service delivery points. Note that the vaccine preventable deaths estimates are vaccine antigen specific, while the excess deaths are dependent on the number of required visits. As vaccination visits group delivery of several vaccines, these have a higher benefit-risk ratio than that for individual antigens.

| Vaccine antigen | Vaccination schedule | Deaths averted by vaccination | Excess COVID-19 deaths | Benefit-risk ratio |
| --- | --- | --- | --- | --- |
| Diphtheria | 6, 10, 14 weeks | 12,944 (10,180-16,539) | 5,674 (846-16,830) | 2 (0-7) |
| Tetanus | 6, 10, 14 weeks | 69,254 (54,268-87,343) | 5,674 (846-16,830) | 12 (2-39) |
| Pertussis | 6, 10, 14 weeks | 271,422 (207,238-344,147) | 5,674 (846-16,830) | 48 (8-155) |
| HepB | 6, 10, 14 weeks | 3,827 (2,578-5,826) | 5,677 (846-16,837) | 1 (0-2) |
| Hib | 6, 10, 14 weeks | 54,840 (49,521-61,230) | 5,696 (849-16,896) | 10 (2-30) |
| PCV | 6, 10, 14 weeks | 46,494 (40,002-55,014) | 5,052 (752-14,979) | 9 (2-29) |
| RotaC | 6, 10 weeks | 10,666 (9,578-11,890) | 2,391 (364-7,221) | 4 (1-14) |
| MCV1 | 9 months | 194,388 (181,469-209,379) | 1,896 (228-5,778) | 103 (16-332) |
| RCV | 9 months | 1,147 (738-1,679) | 744 (85-2,264) | 2 (0-5) |
| MenA | 9 months | 460 (335-665) | 280 (34-856) | 2 (0-6) |
| YFV | 9 months | 23,345 (17,426-30,929) | 875 (100-2,664) | 27 (4-87) |
| MCV2 | 15-18 months | 10,282 (9,354-11,237) | 751 (81-2,277) | 14 (2-45) |
| EPI-1<br>(DTP3, HepB3, Hib3,<br>PCV3, RotaC) | 6, 10, 14 weeks | 471,068 (406,088-548,290) | 5,696 (849-16,896) | 82 (14-261) |
| EPI-2<br>(MCV1, RCV1, MenA,<br>YFV) | 9 months | 219,726 (204,572-235,744) | 1,896 (228-5,778) | 116 (18-374) |
| EPI<br>(DTP3, HepB3, Hib3,<br>PCV3, RotaC, MCV1,<br>RCV1, MenA, YFV,<br>MCV2) | 6, 10, 14 weeks; 9<br>months; 15-18 months | 701,828 (635,416-782,050) | 8,341 (1,280-25,029) | 84 (14-267) |

**Table 2.** Benefits and risks of sustaining routine childhood immunisation at the national level in the African countries. The benefit-risk ratio estimates (median estimates and 95% uncertainty intervals) show the child deaths averted by sustaining routine childhood immunisation in the African countries per COVID-19 death attributable to excess SARS-CoV-2 infections acquired through visiting routine vaccination service delivery points. The combined impact of the routine childhood immunisation is shown for 3-dose DTP3, HepB3, Hib3, PCV3 for children at 6, 10 and 14 weeks, 2-dose RotaC for children at 6 and 10 weeks, 1-dose MCV1, RCV1, MenA, YFV for children at 9 months, and 1-dose MCV2 for children at 15-18 months of age.

| Country | Deaths averted by vaccination | Excess COVID-19 deaths | Benefit-risk ratio |
| --- | --- | --- | --- |
| Angola | 26,156 (18,377-36,792) | 146 (24-434) | 180 (28-598) |
| Burundi | 8,640 (5,946-12,784) | 79 (13-238) | 110 (19-367) |
| Benin | 7,285 (4,817-11,246) | 78 (8-228) | 95 (14-311) |
| Burkina Faso | 14,103 (9,626-20,955) | 180 (20-534) | 80 (13-259) |
| Botswana | 989 (646-1,511) | 15 (2-43) | 70 (11-233) |
| Central African Republic | 2,353 (1,585-3,513) | 20 (3-60) | 119 (21-392) |
| Côte d'Ivoire | 19,401 (12,712-28,863) | 194 (20-571) | 102 (16-339) |
| Cameroon | 13,031 (8,735-19,862) | 176 (20-519) | 75 (11-249) |
| Congo - Kinshasa | 61,538 (41,399-92,956) | 563 (88-1,674) | 111 (18-371) |
| Congo - Brazzaville | 3,370 (2,366-4,919) | 21 (3-63) | 160 (19-515) |
| Comoros | 420 (267-661) | 7 (1-22) | 58 (7-197) |
| Cape Verde | 148 (85-251) | 3 (0-9) | 52 (6-176) |
| Djibouti | 273 (173-435) | 5 (1-14) | 58 (8-203) |
| Algeria | 18,164 (11,750-28,146) | 268 (29-794) | 69 (10-234) |
| Egypt | 24,593 (11,655-48,336) | 412 (54-1,221) | 60 (6-216) |
| Eritrea | 2,103 (1,380-3,152) | 29 (4-88) | 74 (9-245) |
| Ethiopia | 60,854 (38,286-95,401) | 866 (100-2,558) | 73 (10-243) |
| Gabon | 866 (554-1,360) | 8 (1-25) | 105 (17-358) |
| Ghana | 18,589 (12,358-26,534) | 219 (32-658) | 86 (14-281) |
| Guinea | 9,307 (6,339-13,372) | 121 (17-362) | 78 (11-255) |
| Gambia | 2,208 (1,560-3,107) | 39 (6-117) | 58 (10-189) |
| Guinea-Bissau | 1,355 (960-1,912) | 12 (1-36) | 113 (18-379) |
| Equatorial Guinea | 360 (231-564) | 4 (0-13) | 83 (12-273) |
| Kenya | 20,030 (12,691-31,736) | 241 (38-720) | 86 (13-292) |
| Liberia | 3,965 (2,913-5,689) | 34 (4-101) | 118 (17-381) |
| Libya | 2,323 (1,517-3,463) | 34 (5-103) | 70 (11-230) |
| Lesotho | 829 (536-1,263) | 15 (2-43) | 57 (8-195) |
| Morocco | 7,273 (3,698-13,837) | 221 (26-657) | 34 (4-124) |
| Madagascar | 14,293 (9,228-22,263) | 136 (21-405) | 107 (16-359) |
| Mali | 13,302 (9,259-18,971) | 144 (17-426) | 94 (14-308) |
| Mozambique | 20,206 (13,487-30,366) | 208 (33-624) | 98 (14-317) |
| Mauritania | 2,720 (1,905-4,119) | 30 (3-90) | 91 (13-310) |
| Mauritius | 260 (177-391) | 4 (1-11) | 71 (11-230) |
| Malawi | 8,923 (5,398-14,737) | 131 (20-393) | 69 (10-232) |
| Namibia | 1,179 (768-1,812) | 19 (2-56) | 63 (9-214) |
| Niger | 21,835 (15,854-30,867) | 262 (30-776) | 85 (13-278) |
| Nigeria | 89,167 (61,172-133,594) | 942 (98-2,773) | 96 (16-316) |
| Rwanda | 8,061 (5,274-12,053) | 87 (14-260) | 94 (15-318) |
| Sudan | 22,338 (13,975-34,110) | 334 (49-1,003) | 68 (10-231) |
| Senegal | 11,306 (7,856-15,866) | 250 (37-758) | 46 (6-154) |
| Sierra Leone | 6,891 (5,096-9,376) | 86 (11-256) | 81 (12-266) |
| Somalia | 9,697 (6,695-14,275) | 102 (11-300) | 96 (16-319) |
| South Sudan | 3,176 (1,716-5,251) | 34 (5-102) | 93 (11-322) |
| São Tomé and Príncipe | 120 (72-186) | 1 (0-4) | 91 (12-296) |
| Swaziland | 346 (180-603) | 9 (1-28) | 38 (4-136) |
| Seychelles | 34 (23-50) | 0 (0-1) | 74 (10-245) |
| Chad | 9,016 (5,998-13,984) | 98 (10-288) | 94 (11-310) |
| Togo | 4,933 (3,206-7,819) | 56 (6-164) | 90 (13-307) |
| Tunisia | 1,854 (723-3,626) | 54 (8-163) | 35 (3-128) |
| Tanzania | 36,630 (23,570-56,036) | 584 (87-1,757) | 64 (8-209) |
| Uganda | 20,906 (12,346-34,358) | 246 (39-734) | 87 (12-299) |
| South Africa | 18,844 (12,575-28,607) | 310 (36-920) | 62 (10-209) |
| Zambia | 11,042 (7,453-16,810) | 121 (19-361) | 93 (13-312) |
| Zimbabwe | 7,759 (5,269-11,124) | 94 (14-284) | 83 (12-278) |

We estimate that the three immunisation visits for EPI-1 add 2.4% (0.7 - 7.6) altogether and each immunisation visit of EPI-2 and EPI-3 add 0.8% (0.2 - 2.7) to the probability of excess SARS-CoV-2 infection in the household. As a result, continuation of routine childhood immunisation in Africa may lead to 8,300 (1,300 - 25,000) excess deaths attributable to additional SARS-CoV-2 infections associated with the immunisation visits of children. About 8 (0 - 40) of these are expected to be among the vaccinated children, 9 (0 - 45) among their siblings, 914 (83 - 2,800) among their parents or adult carers, and 7,300 (852 - 22,300) among older adults in the household.

For every one excess COVID-19 death attributable to additional household exposure to SARS-CoV-2 infections due to routine childhood immunisation visits, we estimate that the routine childhood immunisation programme would prevent 84 (14-267) deaths in children until 5 years of age in Africa (see Table 1). The benefit of the three EPI-1 immunisation visits in early infancy and the visit for EPI-2 at 9 months were 82 (14-261) and 116 (18-374) deaths averted among children per excess COVID-19 death, respectively. The incremental benefit of the second dose of measles vaccination during EPI-3 visit at 15-18 months was 14 (2-45) deaths averted among children per excess COVID-19 death. Nearly 90% of the excess COVID-19 risk is due to the high fatality rate among older adults aged above 60 years. If only the risk to vaccinated infants is considered, the benefit-risk ratio is substantially higher at 85,000 (4,900 - 546,000) (see appendix A4). Our findings were largely similar across countries (see Figure 1, Table 2, and appendix A5). Country-specific benefit-risk ratios for EPI-1, EPI-2, and EPI-3 are presented in the appendix (see A6, A7, A8). The overall benefit risk-ratio of sustaining routine childhood immunisation ranged from 34 (3-124) in Morocco to 180 (27-598) in Angola, and the number of child deaths averted through vaccination substantially exceeded the number of excess COVID-19 deaths for all the 54 countries of Africa.

**Figure 1.**
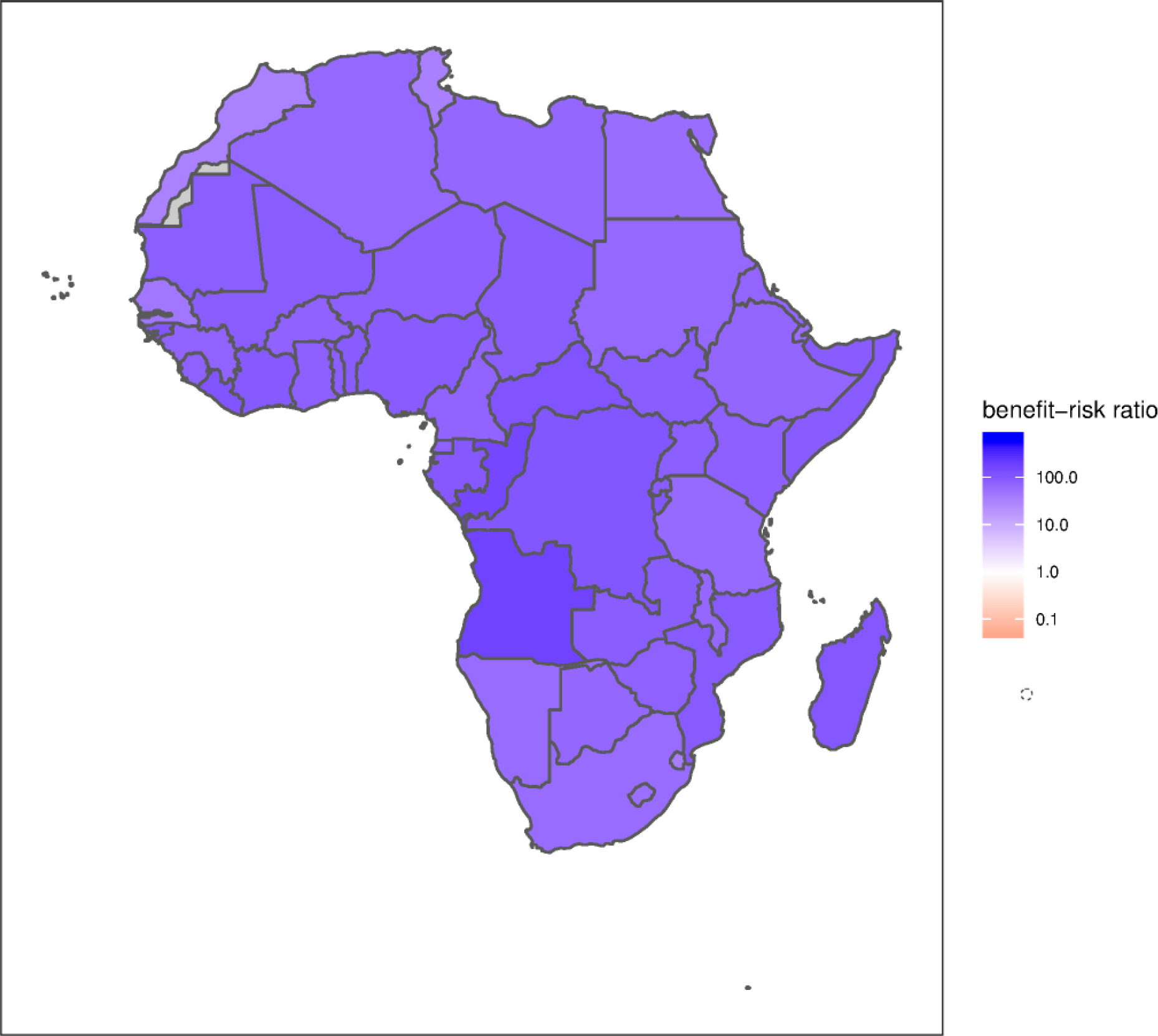
Benefit-risk ratios for sustaining routine childhood immunisation during the COVID-19 pandemic in the African countries. The number of vaccine preventable future deaths averted before completing five years of age by sustaining routine childhood vaccination of DTP, HepB, Hib, PCV, RotaC, MCV, RCV, MenA and YFV per COVID-19 death attributable to excess SARS-CoV-2 infections acquired through visiting routine vaccination service delivery points. The routine childhood vaccines considered are 3-dose DTP3, HepB3, Hib3, PCV3 for children at 6, 10 and 14 weeks, 2-dose RotaC for children at 6 and 10 weeks, 1-dose MCV1, RCV1, MenA, YFV for children at 9 months, and 1-dose MCV2 for children at 15-18 months of age. A benefit-risk ratio larger than 1 indicates in favour of sustaining the routine childhood immunisation programme during the COVID-19 pandemic. * Grey colour indicates missing data. We take a neutral position with respect to territorial claims in the maps.

In the low-impact scenario that approximates the health benefits to only the child deaths averted from measles outbreaks, the benefit-risk ratio to the households of vaccinated children is 3 (0 - 10). When the risk to only the vaccinated children is considered, the benefit-risk ratio is 3,000 (182 - 21,000). Even under these highly conservative assumptions, the benefit ratios for most countries in Africa are larger than 1 and indicates in favour of sustaining the routine childhood immunisation programme during the COVID-19 pandemic (see Figure 2). Tunisia, Eswatini, Morocco, and Egypt have benefit-risk ratios lower than 1, since measles vaccination impact is relatively at the lower end in these three countries in comparison to other countries in Africa. While the lower bounds (of the credible intervals) of the benefit-risk ratios at the household level are lower than 1 for some countries, the lower bounds of the benefit-risk ratios for the vaccinated children are greater than 1 for all countries.

**Figure 2.**
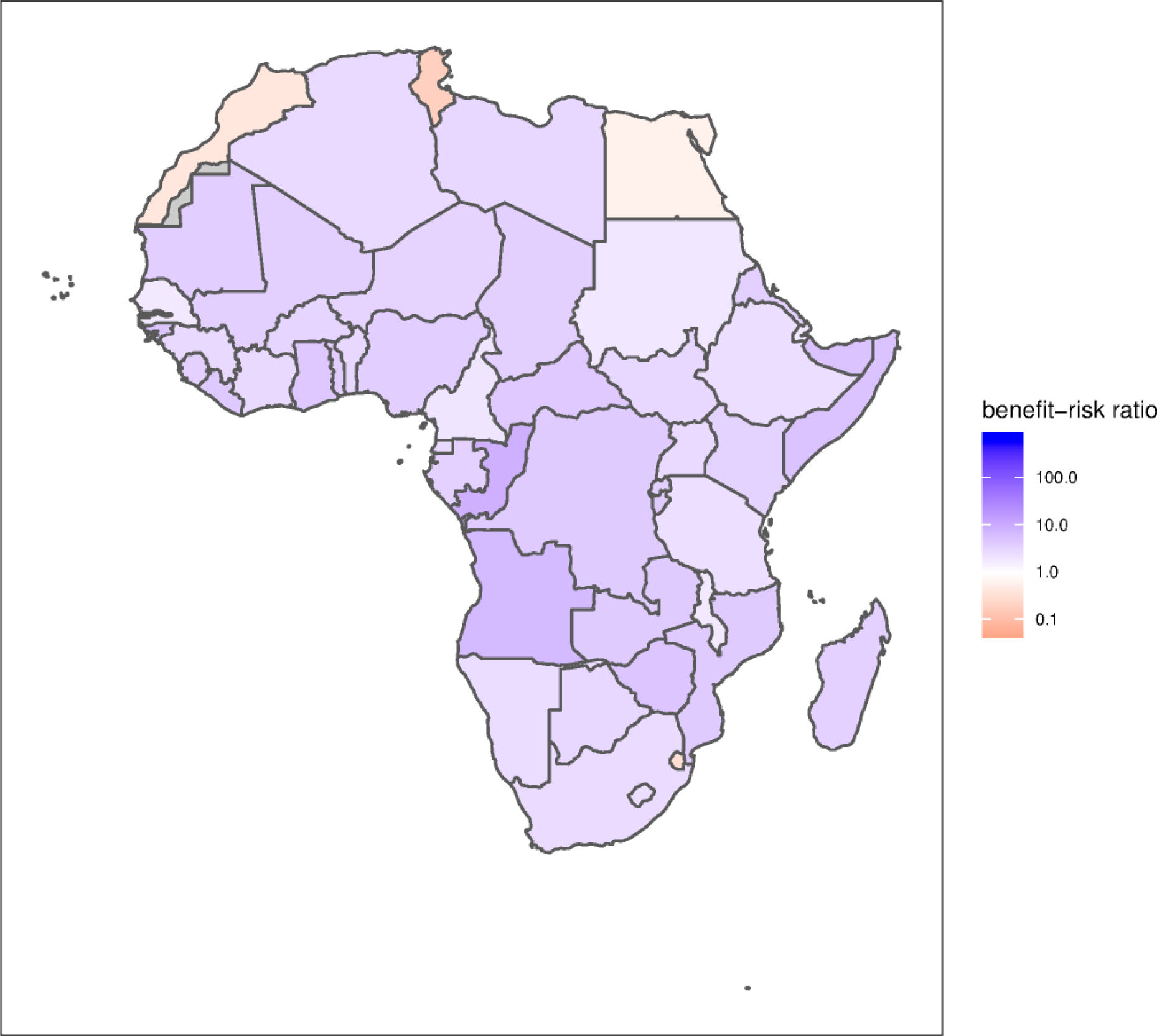
Scenario of measles-only vaccination impact during the COVID-19 pandemic. The number of vaccine preventable future deaths averted before completing five years of age by sustaining routine childhood vaccination of DTP, HepB, Hib, PCV, RotaC, MCV, RCV, MenA and YFV per COVID-19 death attributable to excess SARS-CoV-2 infections acquired through visiting routine vaccination service delivery points. We consider a small chance (12.5%) of measles outbreaks while no other vaccine preventable disease outbreaks take place due to herd immunity. * Grey colour indicates missing data. We take a neutral position with respect to territorial claims in the maps.

We evaluated the contribution of the uncertainty in the model parameters to the uncertainty in the benefit-risk ratio estimates (Figure 3). The main factors influencing our estimates of the benefit-risk ratio were the average number of contacts of the child and their carer during a visit to the vaccine clinic, the average number of transmission relevant contacts of a community member per day and hence the risk for transmission given a potentially infectious contact, and the infection-fatality rate for SARS-CoV-2 infected older adults aged 60 years and above.

**Figure 3.**
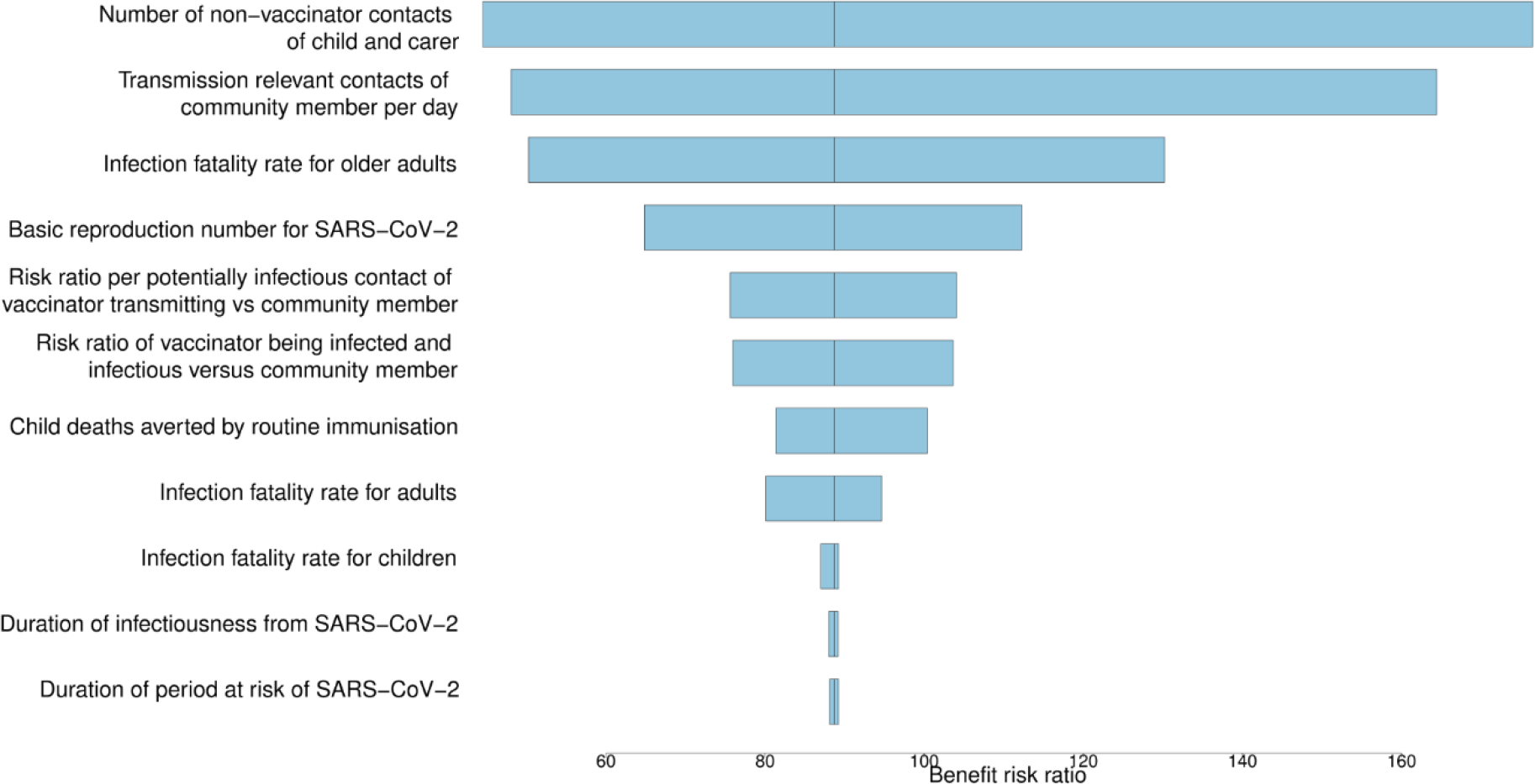
Sensitivity analysis for attributable impact of model parameters on uncertainty in the benefit-risk ratio estimates. Sensitivity analysis shows the estimated contribution of different model parameters to the overall uncertainty in the benefit-risk ratio of sustaining routine childhood immunisation during the COVID-19 pandemic in Africa. The tornado diagram was constructed using a multivariate Poisson regression model to the estimated posterior distribution of the benefit-risk ratio using our model input parameters as predictors, and treating total deaths averted by childhood immunisation as a single variable. The main factors influencing the benefit-risk ratio estimates were the average number of contacts of the child and their carer during a visit to the vaccination clinic, the average number of transmission relevant contacts of a community member per day, and the infection-fatality rate of SARS-CoV-2 infected older adults aged above 60 years.

## Discussion

Our analysis suggests that the benefit from sustaining routine childhood immunisation in Africa far outweighs the excess risk of COVID-19 deaths due to the additional risk for SARS-CoV-2 infections during the child’s vaccination visit, particularly for the vaccinated children. This reinforces the guidance and statement issued by the World Health Organization and the Measles & Rubella Initiative respectively to sustain routine childhood immunisation programmes where essential health services have operational capacity of adequate human resources and vaccine supply while maintaining physical distancing and other infection control measures to ensure the safety of communities and health workers.^15,30^

We base our analyses on model-based country and antigen-specific vaccine impact estimates in low and middle income countries for 2020.^3,18^ There is considerable uncertainty in the impact of suspending immunisation activities for a period of about 6 months and whether a timely and high-coverage catch-up campaign can be conducted soon after. Therefore, we presented two extreme scenarios – high-impact and low-impact, for the potential benefits from sustaining routine childhood vaccination.

In the high-impact scenario, we approximate the impact of sustained routine childhood immunisation with the estimates of vaccination impact for a 6-month cohort in 2020. This is a basic estimate of the likely impact which is also governed by herd protection and physical distancing. While pathogen resurgence will happen gradually due to herd protection from the rest of the population and potentially only once physical distancing is lifted, this could be counterbalanced by unvaccinated children of this and other cohorts continuing to be at risk of disease beyond the 6-month window. In the presence of physical distancing measures, the exposure to non-coronavirus pathogens will also likely be reduced but those who may remain susceptible as a result of immunisation service suspension may get infected once physical distancing measures are relaxed. In the low-impact scenario, we approximate the impact of sustaining vaccination by the number of child deaths as a result of potential measles outbreaks during the COVID-19 risk period while also accounting for catch-up campaigns to be delivered at the end of the COVID-19 risk period. We show that in both scenarios that continuation of routine childhood immunisation is beneficial and outweighs the excess risk of COVID-19 deaths due to the additional risk for SARS-CoV-2 infections during the immunisation visits, especially for the vaccinated children. While the health benefits of routine vaccination are highly beneficial to the children, infections acquired by the children and/or their adult carers and parents at the vaccination clinics pose a risk primarily for the older adults in the households of the vaccinated children. This highlights the value of shielding older adults to lower their risk of acquiring SARS-CoV-2 infections while children in their households can benefit from routine vaccination to lower their risk of acquiring vaccine-preventable infectious diseases.^31^

While the younger African age-demographic may mitigate some of the COVID-19 disease burden, infection fatality rates in Africa may be substantially higher because of the prevalence of likely risk factors including HIV, tuberculosis, and malnutrition as well as lack of access to antibiotics to limit the risk for bacterial coinfections in some parts of Africa. In the event that infection fatality rates in Africa turn out to be higher than elsewhere, then the estimated benefit-risk ratio would be reduced. However, our uncertainty analysis illustrates that while the uncertainty of the COVID-19 infection fatality rate is a key factor in the overall uncertainty of our estimates, even at the upper mortality bounds, sustaining routine childhood vaccination is beneficial. Furthermore, the effects of a potentially higher COVID-19 case-fatality ratio in Africa may be balanced by a higher case-fatality ratio of measles and the other vaccine preventable diseases in times when the healthcare system is stretched, treatment supplies are disrupted, and access to drugs such as vitamin A and antibiotics are limited.

Our findings were similar across countries with respect to the benefit-risk ratios indicating in favour of sustaining the childhood immunisation programmes during the COVID-19 pandemic in Africa. Although there will be heterogeneity in implementation and compliance of prevention and control measures for COVID-19 among the different countries, the benefits of sustaining childhood immunisation far outweigh the risks of excess SARS-CoV-2 infections acquired during the vaccination visits, especially for the vaccinated children.

Because of high transmissibility of measles, routine childhood immunisation coverage in many countries is insufficient to prevent outbreaks. To aid routine vaccination coverage, supplementary immunisation activities are conducted regularly, many of them scheduled for this year, at a point shortly before sufficient population immunity has built up to prevent measles outbreaks.^32^ Many supplementary immunisation activities have recently been postponed to reduce the risk for COVID-19 infections during mass vaccination,^15^ further enhancing the likelihood and impact of measles outbreaks if routine childhood vaccination is suspended. Since they are timed at the right intervals to avoid outbreaks, our low-impact scenario is likely to underestimate the risk of an outbreak occurring due to suspension of supplementary immunisation activities. While this may in part be mitigated by reduced contact patterns in response to COVID-19, the risk of outbreaks will be concentrated in the periods where interventions are gradually lifted and before a campaign can be conducted.

We conducted a probabilistic sensitivity analysis to assess the impact of parameter uncertainty on the estimated benefit-risk ratios. We found that the biggest contribution to the uncertainty around the benefit of sustaining routine childhood immunisation during the COVID-19 pandemic in Africa are the transmission probability and the number of contacts during a vaccination visit. This highlights the need for personal protective equipment for vaccinators, the need to implement physical distancing measures including the avoidance of crowded waiting rooms for vaccination visits, and the importance of good hygiene practices to reduce the risk of SARS-CoV-2 acquisition and transmission at the vaccination clinics. While It will be challenging to implement some of these infection prevention and control measures in many African countries due to resource constraints, the risks can be minimised and the benefits can be enhanced by providing immunisation bundled with other health services, thereby reducing the number of physical touch points with the health system.

We estimated the benefit-risk trade-off for sustaining routine childhood immunisation during the COVID-19 pandemic in Africa and found that the benefits substantially outweigh the risks. However, there are other factors that must be considered for strategic decision making to sustain routine childhood immunisation in African countries during the COVID-19 pandemic. These include logistical constraints of vaccine supply and delivery cold chain problems caused by the COVID-19 pandemic, reallocation of doctors and nurses to other prioritised health services, healthcare staff shortages caused by SARS-CoV-2 infections among the staff from symptomatic and asymptomatic infectious individuals or staff shortages because of ill-health or underlying health conditions that put them at increased risk for severe COVID-19 disease, and decreased demand for vaccination arising from community reluctance to visit vaccination clinics for fear of contracting SARS-CoV-2 infections as well as broader distrust of vaccines fueled by COVID-19 related rumours. Also, the opportunity risk of SARS-CoV-2 infection for the vaccinated children and healthcare staff involved in immunisation activities as well as to their households and onward SARS-CoV-2 transmission into the wider community should be considered (see appendix A9).

While we have estimated benefit-risk ratios based on deaths averted, the analysis can be extended to estimate benefit-risk ratios based on disability-adjusted life-years averted or quality-adjusted life-years gained. Since the deaths averted by vaccination are primarily among under-5-year-old children and deaths caused by COVID-19 are primarily among older adults, the benefit-risk ratios will be relatively higher using DALYs or QALYs and more favourable towards sustaining routine childhood immunisation programmes during the COVID-19 pandemic in Africa.

In conclusion, routine childhood immunisation programmes should be safeguarded for continued service delivery and prioritised for the prevention of infectious diseases, as logistically possible, as part of delivering essential health services during the COVID-19 pandemic in Africa.

## Data Availability

The program code and data for the benefit-risk analysis conducted in this study is accessible on GitHub (https://github.com/vaccine-impact/epi_covid).

## Acknowledgements

We thank Nicholas Grassly, Raymond Hutubessy, and Anthony Scott for helpful discussions. We thank the reviewers for their valuable feedback on previous versions of the manuscript.

This study was supported in part by Gavi, the Vaccine Alliance (KA, AC, MJ, PK); Bill & Melinda Gates Foundation (OPP1157270: KA, AC, MJ, PK; INV-003174: MJ); Vaccine Impact Modelling Consortium (KA, AC, MJ, PK); Elrha R2HC/UK DFID (KvZ); UK Department of Health and Social Care (16/137/109: MJ; Health Protection Research Unit for Modelling Methodology HPRU-2012-10096: NGD; UK Public Health Rapid Support Team: TJ); HDR UK (MR/S003975/1: RME); UK MRC (MC_PC 19065: RME; MR/P014658/1: GMK); Alan Turing Institute (AE); DFID/Wellcome Trust (Epidemic Preparedness Coronavirus research programme 221303/Z/20/Z: CABP, KvZ); ERC Starting Grant (#757688: CJVA, KEA; #757699: JCE, RMGJH); European Union’s Horizon 2020 research and innovation programme - project EpiPose (101003688: KP, PK, WJE, YL); RCUK and ESRC (ES/P010873/1: AG, CIJ, TJ); The Nakajima Foundation (AE); NIHR (16/137/109: BJQ, CD, FYS, YL); Health Protection Research Unit for Modelling Methodology HPRU-2012-10096: TJ; PR-OD-1017-20002: AR); Royal Society (RP\EA\180004: PK); UK DHSC/UK Aid/NIHR (ITCRZ 03010: HPG); and Wellcome Trust (206250/Z/17/Z: AJK, TWR; 208812/Z/17/Z: SC, SFl; 210758/Z/18/Z: JDM, JH, NIB, SA, SFu).

## Contributors

KA, SRP, MJ and SFl conceptualised the study and wrote the original draft; KA, SRP, KvZ, AC, SFu, TM, DH, ED, MJ and SFl developed the benefit-risk model; KA compiled the data sets; KA and SRP developed the software and conducted the analysis, and all authors contributed to reviewing and editing of the manuscript and have approved the final version.

## Supplementary appendix

A1. Simulation parameters for SARS-CoV-2 infection dynamics

A2. COVID-19 risk model

A3. Household structure, age composition and infection fatality risk

A4. Age and antigen specific benefit-risk ratios for Africa at the continental level

A5. Country and age specific benefit-risk ratios for Africa at the national level

A6. Benefit-risk ratio of vaccines delivered in the first, second, and third vaccination-related clinical visits

A7. Benefit-risk ratio of vaccines delivered in the fourth vaccination-related clinical visit

A8. Benefit-risk ratio of vaccines delivered in the fifth vaccination-related clinical visit

A9. Opportunity risk for vaccinated children and healthcare staff involved in immunisation activities

A10. Age and antigen specific deaths averted by vaccination, excess deaths due to COVID-19, and benefit-risk ratios for Africa at the continental level

A11. Country, age, and antigen specific deaths averted by vaccination, excess deaths due to COVID-19, and benefit-risk ratios for Africa at the national level

A12. Age and antigen specific deaths averted by measles vaccination, excess deaths due to COVID-19, and benefit-risk ratios for Africa at the continental level – Scenario of measles-only vaccination impact

A13. Country and age specific deaths averted by measles vaccination, excess deaths due to COVID-19, and benefit-risk ratios for Africa at the national level – Scenario of measles-only vaccination impact

## A1. Simulation parameters for SARS-CoV-2 infection dynamics

**Table A1.**
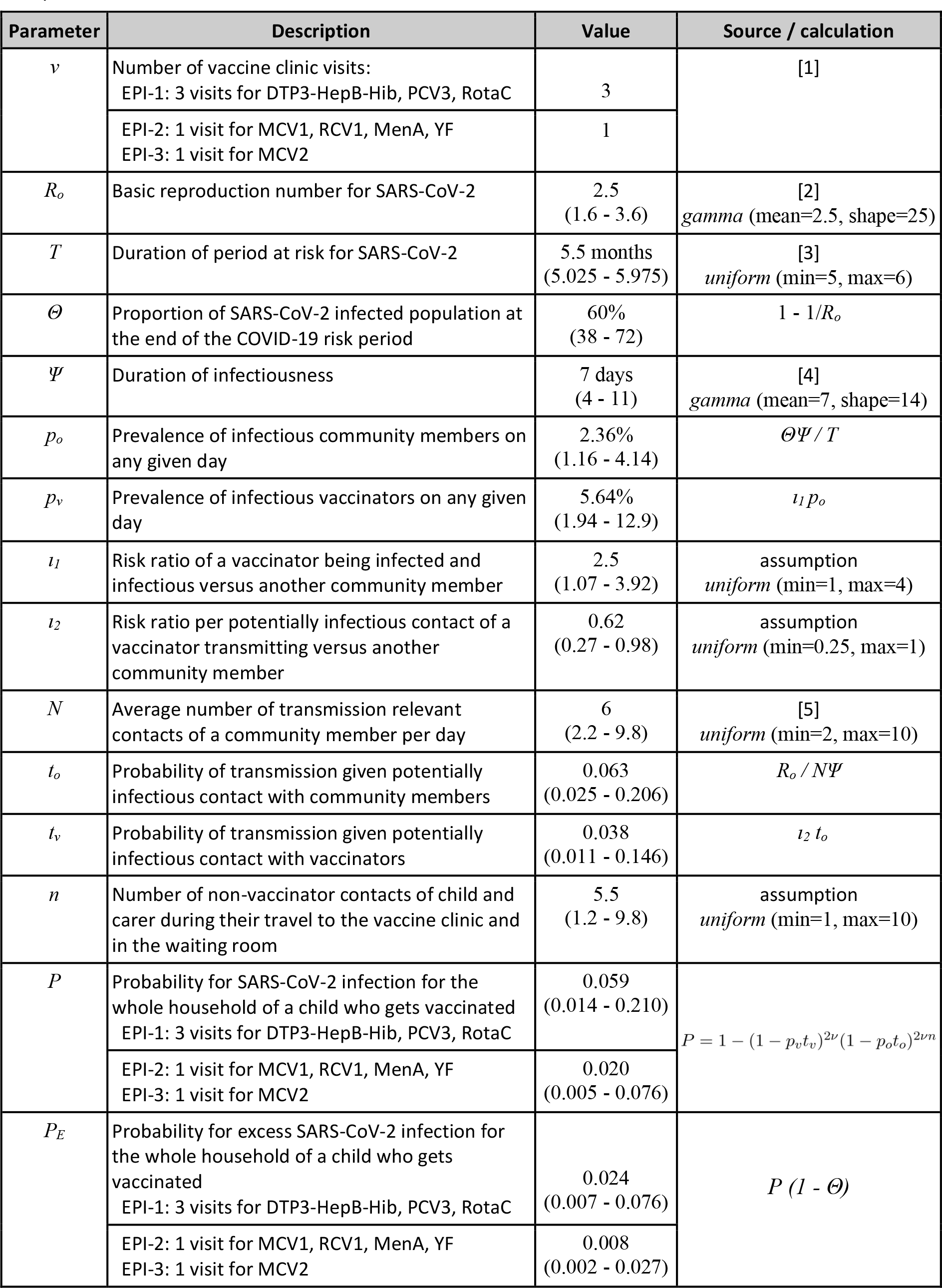
Parameters governing the estimation of SARS-CoV-2 infection dynamics during immunisation visits – baseline values & 95% uncertainty intervals for probabilistic sensitivity analyses.

## A2. COVID-19 risk model

The risk of infection with SARS-CoV-2 depends on the stage of the epidemic, with relatively higher risk during the early incline phase of the epidemic and larger proportion of susceptible population and relatively lower risk during the late decline phase of the epidemic and smaller proportion of susceptible population. We refer excess risk to additional infections among households that are attributable to the vaccination visits, that these additional infections among household members would not have occurred during the course of the epidemic if not for the vaccination visits.

As a base case, we assume that through contact reducing interventions, community SARS-CoV-2 transmission will be spread over a period (*T*) of 5 to 6 months and the exposure risk is constant during that time due to contact-reducing interventions successfully mitigating sharp peaks in disease (Table A1) [3]. We assume that these measures will be gradually lifted and that, in the absence of vaccination visits, between *Θ = 40%* and *Θ = 70%* of the population will have been infected with SARS-CoV-2. This corresponds for example to the herd immunity threshold for a basic reproduction number (*R*_*o*_) of between 1.6 and 3.6 assuming that everyone who is infected develops full immunity. Partial immunity following infection combined with a reduction in effective reproduction number following physical distancing measures would also achieve a final epidemic size of around this level. It follows from above that between 30% and 60% of the population would not have become infected with SARS-CoV-2 independent of whether or not the infants in their households had attended routine childhood vaccination. Furthermore, if after 6 months 60% of the population was infected then, assuming a duration of infectiousness (*Ψ*) of one week [4] and a reasonably flat epidemic curve, then on any given day about *p*_*o*_ ∼ *2%* of the population would be infected and potentially transmitting. In comparison to community members, we assume that vaccinators are at higher risk of being infected (between 1 and 4 times, *p*_*v*_ = *ι*_*1*_ *p*_*o*_) because of their higher frequency of exposure to other people, but at lower risk of onward transmission (between 0.25 and 1 times, *t*_*v*_ = *ι*_*2*_ *t*_*o*_) because most of their contacts with vaccinees are brief, and they have enhanced risk awareness and use corresponding protective measures including basic respiratory hygiene and personal protective equipment as available. Also, we assume that an infant child and the parent or adult carer each have between 1 and 10 (*n = U*(*1, 10*)) potentially infectious contacts during their travel to the vaccine clinic and in the waiting room.

For each of the potentially infectious contacts by the child and parent with community members, there is a probability of transmission (*t*_*o*_ *= R*_*o*_ */ NΨ*), which for example corresponds to (*t*_*o*_ ∼ *6%*) probability of a transmission event occurring for (*R*_*o*_ *= 2*.*5*) secondary infections for someone with 6 contacts per day during their infectious period of 7 days (i.e., a community member) or 21 potentially infectious contacts per day but who self isolates on symptom onset that occurred 2 days into their infectious period (i.e., a vaccinator).

Both the vaccinated child and the parent or caregiver, will be at additional risk of exposure during travel to the vaccine clinic, while waiting at the vaccine clinic and during vaccination. In addition, we assume that if either of them gets infected they will infect all other household members, owing to the high secondary attack rates observed for family gatherings [6]. We ignore any additional secondary infections outside the household, which are likely to be minimal due to physical distancing measures.

Based of the Reed-Frost epidemic model [7], the probability (*P*) for a SARS-CoV-2 infection for the whole household of a child who gets vaccinated is calculated as one minus the probability of either the infant or the mother not being infected by either the vaccinator or anyone else on any of the vaccination visits: *P* = 1 − (1 − *p*_*v*_*t*_*v*)_^2*ν*^ (1 − *p*_*o*_*t*_*o*_)^2*νn*^ with *v* the number of vaccine clinic visits. Hence, the probability for such infection to be in excess of SARS-CoV-2 infections that would have occurred otherwise is *P*_*E*_ *= P* (*1 - Θ*).

We assume that during the 6 months of SARS-CoV-2 transmission, all children who get one dose of DTP will also get the other two doses. However, children receiving their measles containing vaccines will only get one dose during that time window because the two doses are given more than six months apart. The number of children who would normally get DTP during the considered time frame is approximated by half of the under one-year old population. Similarly, the number of children who will get either the first or the second measles-containing vaccine dose is half of the under 1-year old children or half of the children aged 12-23 months respectively.

## A3. Household structure, age composition and infection fatality risk

We use the country-specific household age composition to approximate the age distribution in households at risk of SARS-CoV-2 infection given that one of the household members is a child who has been vaccinated [8]. First, we estimate the number of siblings of an infant from the average number of household members aged less than 20 in households with at least one member aged less than 20. The number of siblings is adjusted to account for the effect of birth order by assuming that on average the infant would be the mid-born child. Secondly, we assume the average household will have two adults (parents or caregivers). Thirdly we assume that a proportion of households with vaccinated children will also have 2 older adults aged over 60 years. We estimate this proportion using the percentage of households that have both members aged less than 20 years and over 60 years old.

To estimate the number of COVID-19 deaths in infected households, we applied age-stratified infection fatality risk for SARS-CoV-2 using estimates obtained from reported cases and their severity in China in combination with the proportion of asymptomatic infections estimated among international residents repatriated from China [9]. For children, we used the reported risks for ages 0-9 years, for adults the risk for ages 30-39 years, and for older adults the risk for ages 60 years and above. To account for uncertainty in these estimates we used gamma distributions fitted to the reported uncertainty in these risks.

**Table A3.** Infection fatality risk parameters used in sensitivity analysis to estimate the number of COVID-19 deaths amongst infected households, based on model-based analysis estimates of the severity of COVID-19 [9].

| Infection fatality rate | Probability distribution | Value |
| --- | --- | --- |
| Children<br>(aged < 20 years) | *gamma* (shape=1.716739, rate=867.9138) | 0.00161%<br>(0.00019 - 0.00586) |
| Adults<br>(aged 20 - 59 years) | *gamma* (shape=8.550388, rate=97.24091) | 0.08464%<br>(0.03926 - 0.15629) |
| Older adults<br>(aged ≥ 60 years) | *gamma* (shape=12.18207, rate=3.607364) | 3.28379%<br>(1.73737 - 5.53980) |

## A4. Age and antigen specific benefit-risk ratios for Africa at the continental level

**Table A4.** Age-and antigen-specific benefit-risk ratios for childhood vaccination during the COVID-19 pandemic in Africa at the continental level. The benefit-risk ratio estimates (central estimates and uncertainty intervals) show the child deaths averted by continuing the routine childhood immunisation programmes per excess COVID-19 death caused by SARS-CoV-2 infections acquired in the vaccination service delivery points in Africa. The routine childhood vaccines considered are 3-dose DTP3, HepB3, Hib3, PCV3 for children at 6, 10 and 14 weeks, 2-dose RotaC for children at 6 and 10 weeks, 1-dose MCV1, RCV1, MenA, YFV for children at 9 months, and 1-dose MCV2 for children at 15-18 months of age. Benefit-risk ratio above 1 indicates in favour of sustaining the routine childhood immunisation programme during the COVID-19 pandemic. The health benefits are accrued by the vaccinated children while the excess COVID-19 risk is disaggregated across the different age groups in the household.

| Vaccine | Benefit-risk ratios |  |  |  |  |
| --- | --- | --- | --- | --- | --- |
|  | Household | Vaccinated children | Siblings<br>(< 20 years of age) | Adults<br>(20-60 years of age) | Older adults<br>(> 60 years of age) |
| Diphtheria (DTP3) | 2 (0-7) | 2,293 (139-14,926) | 2,014 (122-13,107) | 21 (4-72) | 3 (0-9) |
| HepB3 | 1 (0-2) | 686 (33-4,688) | 602 (29-4,116) | 6 (1-22) | 1 (0-3) |
| Hib3 | 10 (2-30) | 9,796 (724-61,874) | 8,593 (635-54,275) | 88 (14-299) | 11 (2-35) |
| MCV1 | 103 (16-332) | 103,318 (6,602-675,385) | 90,700 (5,796-592,902) | 934 (147-3,218) | 117 (17-384) |
| MCV2 | 14 (2-45) | 13,620 (708-87,942) | 13,107 (682-84,629) | 123 (17-425) | 16 (2-51) |
| PCV3 | 9 (2-29) | 9,489 (706-61,053) | 8,210 (611-52,826) | 86 (14-294) | 10 (2-34) |
| Pertussis (DTP3) | 48 (8-155) | 47,980 (2,858-320,376) | 42,132 (2,510-281,330) | 441 (70-1,509) | 54 (10-179) |
| RCV1 | 2 (0-5) | 1,601 (82-10,670) | 1,417 (72-9,441) | 14 (2-51) | 2 (0-6) |
| RotaC | 4 (1-14) | 4,732 (258-30,782) | 4,241 (232-27,583) | 43 (7-145) | 5 (1-16) |
| Tetanus (DTP3) | 12 (2-39) | 12,436 (728-80,478) | 10,920 (639-70,670) | 111 (14-385) | 14 (2-46) |
| YFV | 27 (4-87) | 27,616 (1,747-184,358) | 21,749 (1,376-145,190) | 248 (32-849) | 31 (4-102) |
| MenA | 2 (0-6) | 1,750 (94-11,681) | 1,355 (73-9,041) | 16 (2-56) | 2 (0-6) |
| DTP3, HepB3, Hib3, PCV3, RotaC | 82 (14-261) | 83,947 (5,017-542,744) | 73,638 (4,401-476,092) | 762 (107-2,595) | 94 (16-303) |
| MCV1, RCV1, MenA, YFV | 116 (18-374) | 116,813 (6,428-759,071) | 102,547 (5,643-666,367) | 1,055 (142-3,615) | 132 (20-431) |
| DTP3, HepB3, Hib3, PCV3, RotaC, MCV1, RCV1, MenA, YFV, MCV2 | 84 (14-267) | 84,674 (4,948-546,123) | 74,892 (4,378-483,020) | 769 (148-2,679) | 96 (14-307) |

## A5. Country and age specific benefit-risk ratios for Africa at the national level

**Table A5.** Country and age specific benefit-risk ratios of vaccines delivered in the five vaccination-related clinical visits (3-dose DTP3, HepB3, Hib3, PCV3; 2-dose RotaC; 1-dose MCV1, RCV1, MenA, YFV, MCV2) during the COVID-19 pandemic in Africa at the country level. The benefit-risk ratio estimates (central estimates and uncertainty intervals) show the child deaths averted by continuing the routine childhood immunisation programmes per excess COVID-19 death caused by SARS-CoV-2 infections acquired in the vaccination service delivery points in Africa. The routine childhood vaccines considered are 3-dose DTP3, HepB3, Hib3, PCV3 for children at 6, 10 and 14 weeks, 2-dose RotaC for children at 6 and 10 weeks, 1-dose MCV1, RCV1, MenA, YFV for children at 9 months, and 1-dose MCV2 for children at 15-18 months of age. Benefit-risk ratio above 1 indicates in favour of sustaining the routine childhood immunisation programme during the COVID-19 pandemic. The health benefits are accrued by the vaccinated children while the excess COVID-19 risk is disaggregated across the different age groups in the household.

| Country | Benefit-risk ratios |  |  |  |  |
| --- | --- | --- | --- | --- | --- |
|  | Household | Vaccinated children | Siblings<br>(< 20 years of age) | Adults<br>(20-60 years of age) | Older adults<br>(> 60 years of age) |
| Angola | 180 (27-598) | 109,538 (7,310-737,985) | 87,995 (5,872-592,844) | 980 (160-3,539) | 227 (33-768) |
| Burundi | 110 (19-367) | 87,595 (4,436-601,184) | 78,952 (3,998-541,869) | 788 (110-2,836) | 131 (16-438) |
| Benin | 95 (14-311) | 102,469 (7,929-691,613) | 81,029 (6,270-546,905) | 936 (159-3,329) | 107 (13-360) |
| Burkina Faso | 80 (13-259) | 87,158 (4,459-587,520) | 60,767 (3,109-409,621) | 794 (105-2,760) | 90 (15-298) |
| Botswana | 70 (11-233) | 78,334 (4,606-512,881) | 92,693 (5,450-606,895) | 695 (94-2,436) | 79 (11-268) |
| Central African Republic | 119 (21-392) | 96,886 (4,313-642,618) | 75,191 (3,347-498,720) | 874 (117-3,022) | 139 (22-470) |
| Côte d'Ivoire | 101 (16-339) | 110,308 (4,615-755,640) | 93,351 (3,906-639,479) | 999 (119-3,545) | 114 (17-390) |
| Cameroon | 75 (11-249) | 91,086 (4,892-598,300) | 70,761 (3,800-464,792) | 820 (103-2,897) | 84 (14-287) |
| Congo - Kinshasa | 111 (18-371) | 91,517 (5,667-634,575) | 68,305 (4,230-473,625) | 823 (95-2,924) | 130 (20-445) |
| Congo - Brazzaville | 160 (19-515) | 111,185 (7,334-785,032) | 112,994 (7,453-797,803) | 1,006 (164-3,597) | 192 (31-659) |
| Comoros | 58 (7-197) | 84,958 (4,504-575,765) | 73,037 (3,872-494,975) | 772 (113-2,742) | 63 (9-219) |
| Cape Verde | 52 (6-176) | 56,845 (3,012-384,901) | 48,598 (2,575-329,064) | 514 (68-1,917) | 58 (7-202) |
| Djibouti | 58 (8-203) | 63,130 (3,235-431,468) | 53,972 (2,766-368,875) | 581 (59-2,109) | 66 (9-233) |
| Algeria | 69 (10-234) | 74,399 (4,421-511,411) | 63,606 (3,780-437,221) | 679 (108-2,475) | 78 (10-272) |
| Egypt | 60 (6-216) | 40,251 (2,074-302,147) | 55,285 (2,848-414,995) | 370 (39-1,458) | 73 (7-272) |
| Eritrea | 74 (9-245) | 80,033 (4,051-526,855) | 68,423 (3,463-450,424) | 720 (103-2,558) | 83 (9-282) |
| Ethiopia | 73 (10-243) | 86,041 (5,101-592,442) | 84,960 (5,037-584,994) | 794 (101-2,790) | 80 (10-274) |
| Gabon | 105 (17-358) | 91,578 (4,602-617,477) | 84,382 (4,240-568,958) | 816 (95-3,046) | 122 (19-424) |
| Ghana | 86 (14-281) | 87,038 (4,608-590,327) | 103,839 (5,498-704,283) | 784 (119-2,766) | 98 (15-323) |
| Guinea | 78 (11-255) | 128,947 (6,320-855,348) | 88,565 (4,341-587,484) | 1,161 (121-4,055) | 84 (11-279) |
| Gambia | 58 (9-189) | 106,616 (7,317-718,292) | 49,744 (3,414-335,133) | 959 (161-3,361) | 62 (9-204) |
| Guinea-Bissau | 113 (18-379) | 121,631 (8,076-809,856) | 103,986 (6,904-692,371) | 1,107 (146-3,922) | 128 (19-441) |
| Equatorial Guinea | 83 (12-273) | 90,947 (3,556-610,982) | 77,754 (3,040-522,348) | 812 (132-2,944) | 93 (13-319) |
| Kenya | 86 (13-292) | 70,406 (3,349-498,310) | 81,892 (3,895-579,604) | 647 (62-2,322) | 99 (14-347) |
| Liberia | 118 (17-381) | 125,265 (8,995-816,096) | 110,302 (7,921-718,618) | 1,114 (136-3,934) | 133 (22-445) |
| Libya | 70 (11-230) | 75,379 (4,094-517,031) | 64,444 (3,500-442,026) | 684 (108-2,411) | 79 (11-264) |
| Lesotho | 57 (8-195) | 80,512 (5,119-565,273) | 118,291 (7,522-830,517) | 730 (83-2,655) | 63 (9-216) |
| Morocco | 34 (3-124) | 43,385 (2,097-317,753) | 47,309 (2,287-346,492) | 395 (35-1,505) | 38 (3-140) |
| Madagascar | 107 (16-359) | 85,342 (5,320-590,034) | 75,798 (4,725-524,052) | 775 (109-2,773) | 126 (18-437) |
| Mali | 94 (15-308) | 111,820 (6,796-741,419) | 81,387 (4,946-539,632) | 1,012 (116-3,503) | 105 (14-348) |
| Mozambique | 98 (14-317) | 77,341 (4,595-510,862) | 76,661 (4,555-506,369) | 704 (105-2,490) | 116 (14-387) |
| Mauritania | 91 (13-310) | 100,042 (6,578-682,233) | 85,529 (5,623-583,262) | 902 (138-3,236) | 102 (14-355) |
| Mauritius | 71 (11-230) | 76,946 (3,535-495,123) | 65,783 (3,023-423,296) | 686 (97-2,453) | 79 (12-267) |
| Malawi | 69 (9-232) | 62,572 (3,514-434,494) | 63,688 (3,577-442,248) | 565 (61-2,107) | 80 (10-274) |
| Namibia | 63 (9-214) | 82,182 (5,366-542,897) | 81,326 (5,310-537,241) | 744 (75-2,626) | 70 (8-237) |
| Niger | 85 (13-278) | 100,340 (6,153-675,769) | 67,251 (4,124-452,920) | 904 (98-3,104) | 95 (15-317) |
| Nigeria | 96 (16-316) | 98,154 (5,286-662,519) | 82,605 (4,448-557,570) | 889 (137-3,095) | 109 (15-367) |
| Rwanda | 94 (15-318) | 77,057 (4,725-516,657) | 90,199 (5,531-604,775) | 693 (97-2,480) | 110 (15-385) |
| Sudan | 68 (10-231) | 70,708 (4,187-473,187) | 56,043 (3,318-375,045) | 643 (104-2,395) | 77 (10-267) |
| Senegal | 46 (6-154) | 99,845 (5,242-690,829) | 45,351 (2,381-313,781) | 912 (111-3,105) | 49 (7-165) |
| Sierra Leone | 81 (12-266) | 121,448 (5,889-812,623) | 94,585 (4,586-632,885) | 1,100 (144-3,738) | 89 (12-295) |
| Somalia | 97 (16-319) | 106,604 (5,416-715,223) | 91,139 (4,630-611,466) | 954 (118-3,365) | 109 (14-365) |
| South Sudan | 93 (11-322) | 82,765 (4,355-555,572) | 58,938 (3,101-395,633) | 746 (115-2,810) | 109 (12-381) |
| São Tomé and Príncipe | 91 (12-296) | 74,944 (4,055-508,305) | 82,903 (4,486-562,283) | 694 (80-2,395) | 107 (13-357) |
| Swaziland | 38 (4-136) | 49,292 (2,433-349,688) | 38,228 (1,887-271,198) | 457 (55-1,682) | 42 (6-153) |
| Seychelles | 74 (10-245) | 80,446 (4,452-559,539) | 68,776 (3,806-478,367) | 726 (88-2,556) | 84 (13-285) |
| Chad | 93 (11-310) | 89,228 (4,713-573,267) | 56,793 (3,000-364,881) | 803 (102-2,786) | 108 (17-367) |
| Togo | 90 (13-307) | 102,242 (5,844-693,935) | 93,667 (5,354-635,735) | 912 (120-3,340) | 101 (13-346) |
| Tunisia | 35 (3-128) | 37,911 (1,427-278,478) | 32,411 (1,220-238,079) | 349 (35-1,313) | 40 (4-147) |
| Tanzania | 64 (8-209) | 69,202 (4,159-458,084) | 61,021 (3,667-403,929) | 627 (98-2,228) | 72 (10-241) |
| Uganda | 87 (12-299) | 75,924 (3,431-532,919) | 65,470 (2,958-459,538) | 692 (90-2,498) | 101 (13-351) |
| South Africa | 62 (10-209) | 75,957 (4,232-537,132) | 113,320 (6,314-801,349) | 682 (91-2,445) | 69 (10-233) |
| Zambia | 93 (13-312) | 78,942 (4,123-525,938) | 62,676 (3,273-417,567) | 709 (99-2,539) | 109 (14-374) |
| Zimbabwe | 83 (12-278) | 87,567 (4,369-596,551) | 101,826 (5,081-693,691) | 783 (116-2,786) | 95 (13-324) |

## A6. Benefit-risk ratio of vaccines delivered in the first, second, and third vaccination-related clinical visits

**Figure A6.**
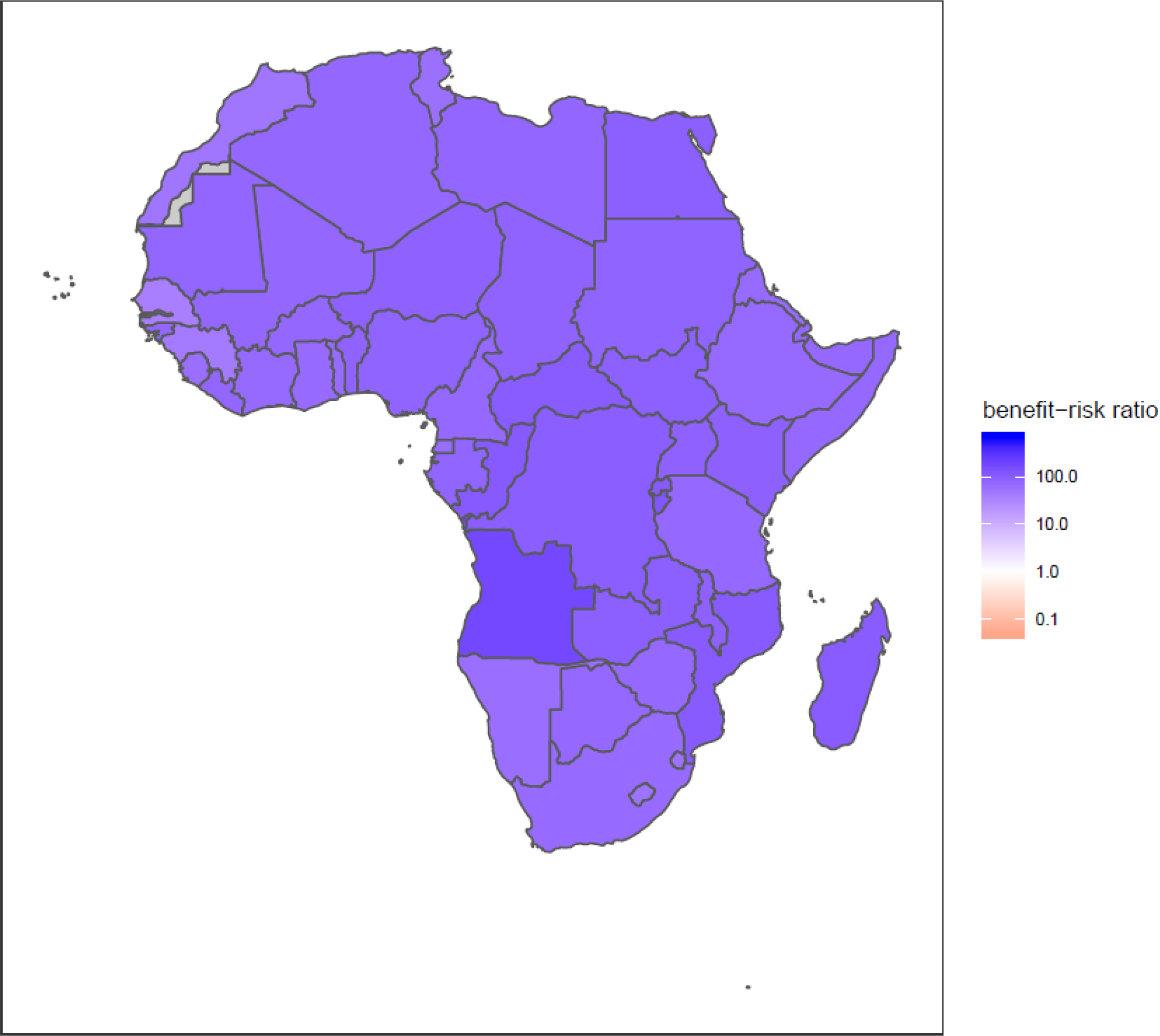
Benefit-risk ratio of vaccines delivered in the first, second, and third vaccination-related clinical visits (3-dose DTP3, HepB3, Hib3, PCV3; 2-dose RotaC) for children at 6, 10, 14 weeks of age during the COVID-19 pandemic in Africa. The central estimates for benefit-risk ratio at the household level show the child deaths averted by continuing the routine childhood immunisation programmes (3-dose DTP3, HepB3, Hib3, PCV3 for children at 6, 10 and 14 weeks of age and 2-dose RotaC for children at 6 and 10 weeks of age) per excess COVID-19 death caused by SARS-CoV-2 infections acquired in the vaccination service delivery points. Benefit-risk ratio above 1 indicates in favour of sustaining the routine childhood immunisation during the COVID-19 pandemic.

## A7. Benefit-risk ratio of vaccines delivered in the fourth vaccination-related clinical visit

**Figure A7.**
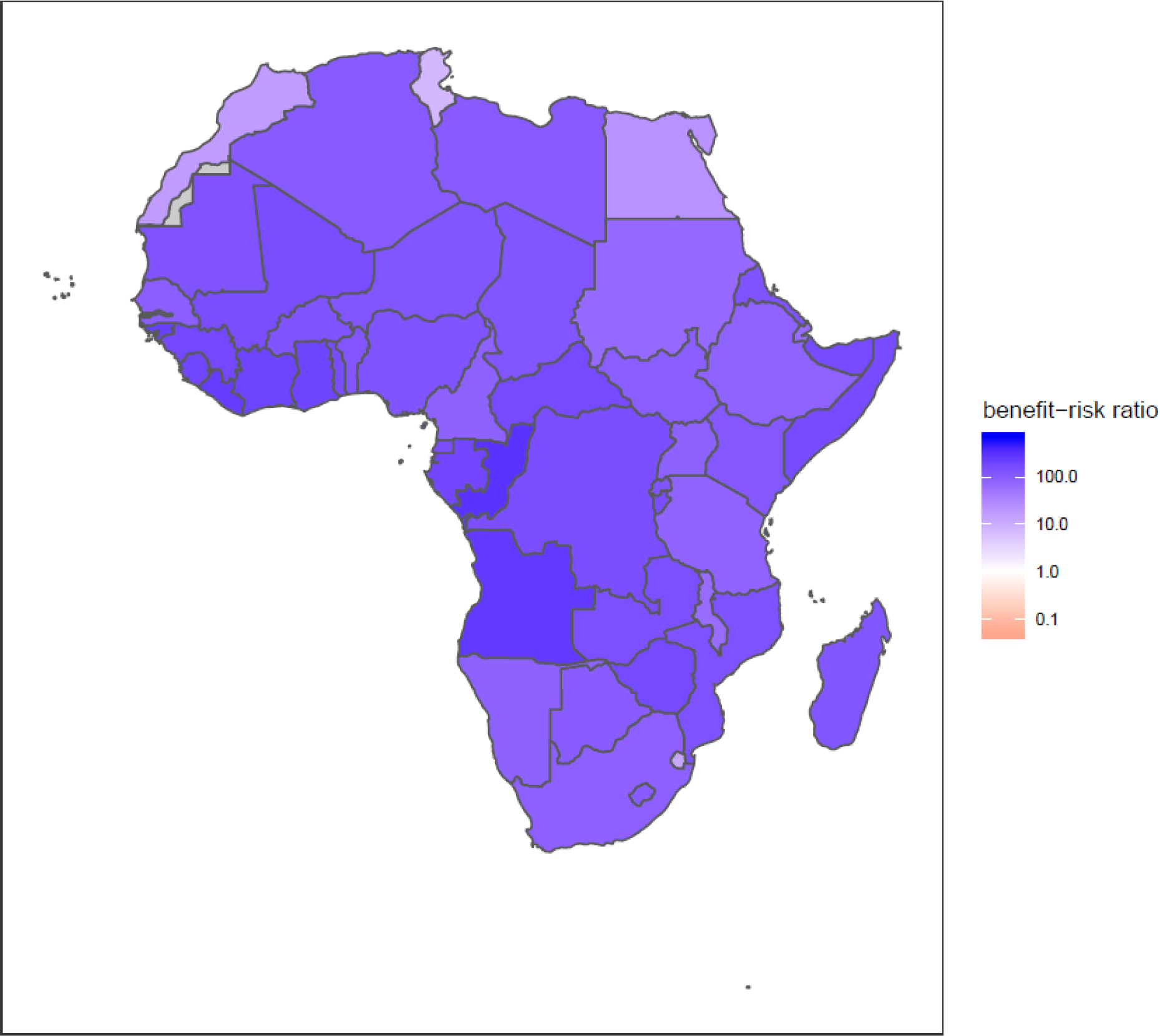
Benefit-risk ratio of vaccines delivered in the fourth vaccination-related clinical visit (1-dose MCV1, RCV1, MenA, YFV) for children at 9-months of age during the COVID-19 pandemic in Africa. The central estimates for benefit-risk ratio at the household level show the child deaths averted by continuing the routine childhood immunisation programmes (1-dose MCV1, RCV1, MenA, YFV for 9-month-old children) per excess COVID-19 death caused by SARS-CoV-2 infections acquired in the vaccination service delivery points. Benefit-risk ratio above 1 indicates in favour of sustaining the routine childhood immunisation during the COVID-19 pandemic.

## A8. Benefit-risk ratio of vaccines delivered in the fifth vaccination-related clinical visit

**Figure A8.**
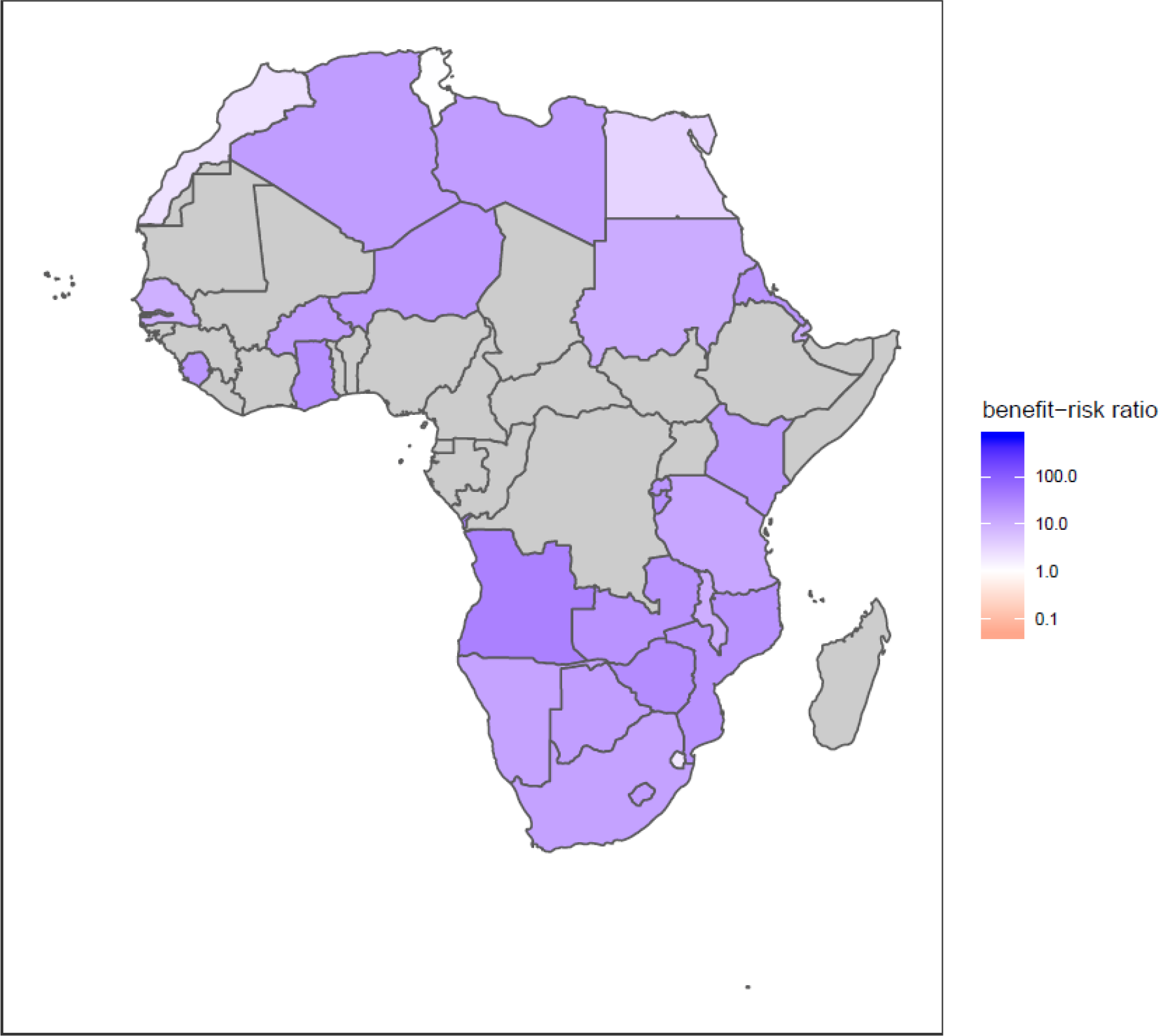
Benefit-risk ratio of vaccines delivered in the fifth vaccination-related clinical visit (1-dose MCV2) for children at 15-18 months of age during the COVID-19 pandemic in Africa. The central estimates for benefit-risk ratio at the household level show the child deaths averted by continuing the routine childhood immunisation programmes (1-dose MCV2 for children aged 15-18 months) per excess COVID-19 death caused by SARS-CoV-2 infections acquired in the vaccination service delivery points. Benefit-risk ratio above 1 indicates in favour of sustaining the routine childhood immunisation during the COVID-19 pandemic. Grey shading indicates null MCV2 coverage.

## A9. Opportunity risk for vaccinated children and healthcare staff involved in immunisation activities

The opportunity risk of SARS-CoV-2 infection for the vaccinated children and healthcare staff involved in immunisation activities as well as to their households and onward SARS-CoV-2 transmission into the wider community should be included in the decision-making process to sustain routine childhood immunisation.

First, we need to know the opportunity risk of SARS-CoV-2 infection for the healthcare staff. Similar to the concept of opportunity cost, what is the risk of SARS-CoV-2 infection to the healthcare staff engaged in alternative healthcare activities if not involved in immunisation activities? If the opportunity risk of alternative healthcare activities is lower than being involved in immunisation activities, then reallocation of healthcare staff from immunisation to alternative healthcare activities is a better risk-avoidance strategy. On the other hand, if the opportunity risk of alternative healthcare activities is higher than being involved in immunisation activities, then healthcare staff face relatively lower risk in continuing to provide the immunisation services, thereby posing relatively lower risk to their households and SARS-CoV-2 transmission into the wider community.

Second, we need to know the opportunity risk of SARS-CoV-2 infection to the vaccinated children. If the alternative activity that the children and their carers would be involved in had a higher risk of SARS-CoV-2 infection in comparison to the risk involved with the immunisation visits, then it is beneficial for the children and their carers to undertake the immunisation visits for the children to get vaccinated and thereby posing relatively lower risk to their households and SARS-CoV-2 transmission into the wider community.

Irrespective of the opportunity risk of SARS-CoV-2 infection for the healthcare staff providing immunisation services during the COVID-19 pandemic, to ensure their safety, health care practices will need to be adapted to minimise the risk of SARS-CoV-2 acquisition and transmission at vaccination clinics. This includes physical distancing measures, personal protective equipment, and good hygiene practices for infection control at the vaccination clinics.

## A10. Age and antigen specific deaths averted by vaccination, excess deaths due to COVID-19, and benefit-risk ratios for Africa at the continental level

Age and antigen specific deaths averted by vaccination, excess deaths due to COVID-19, and benefit-risk ratios (central estimates and uncertainty intervals) for routine childhood vaccination are included in the dataset. The routine childhood vaccines considered are 3-dose DTP3, HepB3, Hib3, PCV3 for children at 6, 10 and 14 weeks, 2-dose RotaC for children at 6 and 10 weeks, 1-dose MCV1, RCV1, MenA, YFV for children at 9 months, and 1-dose MCV2 for children at 15-18 months of age. Note that the risk is disaggregated across the different age groups in the household.

See supplementary appendix 2 (spreadsheet) for the dataset.

## A11. Country, age, and antigen specific deaths averted by vaccination, excess deaths due to COVID-19, and benefit-risk ratios for Africa at the national level

Country, age and antigen specific deaths averted by vaccination, excess deaths due to COVID-19, and benefit-risk ratios (central estimates and uncertainty intervals) for routine childhood vaccination are included in the dataset. The routine childhood vaccines considered are 3-dose DTP3, HepB3, Hib3, PCV3 for children at 6, 10 and 14 weeks, 2-dose RotaC for children at 6 and 10 weeks, 1-dose MCV1, RCV1, MenA, YFV for children at 9 months, and 1-dose MCV2 for children at 15-18 months of age. Note that the risk is disaggregated across the different age groups in the household.

See supplementary appendix 2 (spreadsheet) for the dataset.

## A12. Age and antigen specific deaths averted by measles vaccination, excess deaths due to COVID-19, and benefit-risk ratios for Africa at the continental level – Scenario of measles-only vaccination impact

Age specific deaths averted by measles vaccination, excess deaths due to COVID-19, and benefit-risk ratios (central estimates and uncertainty intervals) for childhood vaccination (measles-only vaccination impact) are included in the dataset. Note that the risk is disaggregated across the different age groups in the household.

See supplementary appendix 2 (spreadsheet) for the dataset.

## A13. Country and age specific deaths averted by measles vaccination, excess deaths due to COVID-19, and benefit-risk ratios for Africa at the national level – Scenario of measles-only vaccination impact

Country and age specific deaths averted by measles vaccination, excess deaths due to COVID-19, and benefit-risk ratios (central estimates and uncertainty intervals) for childhood vaccination (measles-only vaccination impact) are included in the dataset. Note that the risk is disaggregated across the different age groups in the household.

See supplementary appendix 2 (spreadsheet) for the dataset.

## Bibliography

1 Andre FE, Booy R, Bock HL, et al. Vaccination greatly reduces disease, disability, death and inequity worldwide. Bull World Health Organ 2008; 86: 140–6.

2 Ozawa S, Mirelman A, Stack ML, Walker DG, Levine OS. Cost-effectiveness and economic benefits of vaccines in low- and middle-income countries: a systematic review. Vaccine 2012; 31: 96–108.

3 Li X, Mukandavire C, Cucunubá ZM, et al. Estimating the health impact of vaccination against 10 pathogens in 98 low and middle income countries from 2000 to 2030. medRxiv 2019; published online Aug 27. DOI:10.1101/19004358.

4 Berkley S. The Power of Vaccines and How Gavi Has Helped Make the World Healthier: 2019 Lasker-Bloomberg Public Service Award. JAMA 2019; published online Sept 10. DOI:10.1001/jama.2019.13190.

5 Piot P, Larson HJ, O’Brien KL, et al. Immunization: vital progress, unfinished agenda. Nature 2019; 575: 119–29.

6 WHO. Immunization Agenda 2030: A Global Strategy to Leave No One Behind. World Health Organization. 2020. https://www.who.int/immunization/immunization_agenda_2030/en/ (accessed April 4, 2020).

7 WHO UNICEF. WUENIC coverage estimates - Vaccines monitoring system 2019 Global Summary Reference Time Series: DTP3. 2019. https://apps.who.int/immunization_monitoring/globalsummary/timeseries/tswucoveragedtp3.html (accessed April 19, 2020).

8 WHO. Coronavirus disease (COVID-19) Pandemic. 2020. https://www.who.int/emergencies/diseases/novel-coronavirus-2019/ (accessed March 25, 2020).

9 Roberton T, Carter ED, Chou VB, et al. Early estimates of the indirect effects of the COVID-19 pandemic on maternal and child mortality in low-income and middle-income countries: a modelling study. Lancet Glob Health 2020; published online May 12. DOI:10.1016/S2214-109X(20)30229-1.

10 Clark A, Jit M, Warren-Gash C, et al. Global, regional, and national estimates of the population at increased risk of severe COVID-19 due to underlying health conditions in 2020: a modelling study. The Lancet Global Health 2020; published online June. DOI:10.1016/S2214-109X(20)30264-3.

11 Huang C, Wang Y, Li X, et al. Clinical features of patients infected with 2019 novel coronavirus in Wuhan, China. Lancet 2020; 395: 497–506.

12 WHO. Coronavirus disease (COVID-19) Pandemic, World Health Organization. 2020. https://www.who.int/emergencies/diseases/novel-coronavirus-2019 (accessed April 10, 2020).

13 WHO. Coronavirus disease (COVID-2019) Situation Report – 84. 2020; published online April 13. https://www.who.int/docs/default-source/coronaviruse/situation-reports/20200413-sitrep-84-covid-19.pdf (accessed April 14, 2020).

14 Nelson R. COVID-19 disrupts vaccine delivery. Lancet Infect Dis 2020; 20: 546.

15 WHO. Guiding principles for immunization activities during the COVID-19 pandemic. World Health Organization. 2020; published online March 26. https://apps.who.int/iris/bitstream/handle/10665/331590/WHO-2019-nCoV-immunization_services-2020.1-eng.pdf (accessed April 4, 2020).

16 PAHO. The Immunization Program in the Context of the COVID-19 Pandemic. Pan American Health Organization. 2020; published online March 26. https://www.paho.org/en/documents/immunization-program-context-covid-19-pandemic-march-2020 (accessed April 3, 2020).

17 Bwaka A, Bita A, Lingani C, et al. Status of the rollout of the meningococcal serogroup A conjugate vaccine in african meningitis belt countries in 2018. J Infect Dis 2019; 220: S140–7.

18 Feikin D, Flannery B, Hamel M, Stack M, Hansen P. Vaccines for Children in Low- and Middle-Income Countries. In: Black R, Temmerman M, Laxminarayan R, Walker N, eds. Disease Control Priorities (third edition): Volume 2, Reproductive, Maternal, Newborn, and Child Health. Washington DC: World Bank, 2016.

19 United Nations. World Population Prospects 2019, United Nations Department of Economic and Social Affairs Population Division. 2019. https://population.un.org/wpp/ (accessed Nov 18, 2019).

20 WHO. Official country reported vaccination coverage estimates time series, World Health Organization. 2019; published online Dec 10. https://apps.who.int/immunization_monitoring/globalsummary/timeseries/tscoveragebcg.html (accessed April 13, 2020).

21 WHO. WHO Guidelines for Epidemic Preparedness and Response to Measles Outbreaks. 1999. https://www.who.int/csr/resources/publications/measles/whocdscsrisr991.pdf (accessed April 19, 2020).

22 Sudfeld CR, Navar AM, Halsey NA. Effectiveness of measles vaccination and vitamin A treatment. Int J Epidemiol 2010; 39 Suppl 1: i48–55.

23 WHO. Vaccine-preventable diseases: Supplementary Immunization Activities Calendar. 2020. https://www.who.int/immunization/monitoring_surveillance/data/Summary_Measles_SIAs.xls (accessed June 1, 2020).

24 O’Reilly K, Auzenbergs M, Jafari Y, Liu Y, Flasche S, Lowe R. Effective transmission across the globe: the role of climate in COVID-19 mitigation strategies. CMMID Repository. 2020; published online March 26. https://cmmid.github.io/topics/covid19/current-patterns-transmission/role-of-climate.html (accessed April 5, 2020).

25 Martinez-Alvarez M, Jarde A, Usuf E, et al. COVID-19 pandemic in west Africa. Lancet Glob Health 2020; published online April 1. DOI:10.1016/S2214-109X(20)30123-6.

26 Abbey H. An examination of the Reed-Frost theory of epidemics. Hum Biol 1952; 24: 201–33.

27 United Nations, Department of Economic and Social Affairs, Population Division. Database on Household Size and Composition 2019. 2019. https://population.un.org/household (accessed April 1, 2020).

28 Verity R, Okell LC, Dorigatti I, et al. Estimates of the severity of coronavirus disease 2019: a model-based analysis. Lancet Infect Dis 2020; 20: 669–77.

29 R Core Team. R: A language and environment for statistical computing. Vienna, Austria: R Foundation for Statistical Computing, 2019.

30 Measles & Rubella Initiative. More than 117 million children at risk of missing out on measles vaccines, as COVID-19 surges. 2020; published online April 14. https://measlesrubellainitiative.org/measles-news/more-than-117-million-children-at-risk-of-missing-out-on-measles-vaccines-as-covid-19-surges/ (accessed April 19, 2020).

31 van Zandvoort K, Jarvis CI, Pearson C, et al. Response strategies for COVID-19 epidemics in African settings: a mathematical modelling study. medRxiv 2020; published online May 3. DOI:10.1101/2020.04.27.20081711.

32 Measles and Rubella Initiative. Measles and Measles-Rubella Supplementary Immunization Activities (SIA) Schedule, 2020. 2020. https://measlesrubellainitiative.org/resources/sia-schedule/ (accessed April 16, 2020).

## Bibliography (for appendix)

1 WHO. WHO recommendations for routine immunization - summary tables [Internet]. 2019 [cited 1 Apr 2020]. Available: https://www.who.int/immunization/policy/immunization_tables/en/

2 Kucharski AJ, Russell TW, Diamond C, Liu Y, Edmunds J, Funk S, et al. Early dynamics of transmission and control of COVID-19: a mathematical modelling study. Lancet Infect Dis. 2020;20: 553–558. DOI:10.1016/S1473-3099(20)30144-4

3 van Zandvoort K, Jarvis CI, Pearson C, Davies NG, CMMID COVID-19 working group, Russell TW, et al. Response strategies for COVID-19 epidemics in African settings: a mathematical modelling study. medRxiv. 2020; DOI:10.1101/2020.04.27.20081711

4 Woelfel R, Corman VM, Guggemos W, Seilmaier M, Zange S, Mueller MA, et al. Clinical presentation and virological assessment of hospitalized cases of coronavirus disease 2019 in a travel-associated transmission cluster. medRxiv. 2020; DOI:10.1101/2020.03.05.20030502

5 Prem K, Cook AR, Jit M. Projecting social contact matrices in 152 countries using contact surveys and demographic data. PLoS Comput Biol. 2017;13: e1005697. DOI:10.1371/journal.pcbi.1005697

6 Liu Y, Eggo RM, Kucharski AJ. Secondary attack rate and superspreading events for SARS-CoV-2. Lancet. 2020;395: e47. DOI:10.1016/S0140-6736(20)30462-1

7 Abbey H. An examination of the Reed-Frost theory of epidemics. Hum Biol. 1952;24: 201–233.

8 United Nations, Department of Economic and Social Affairs, Population Division. Database on Household Size and Composition 2019 [Internet]. 2019 [cited 1 Apr 2020]. Available: https://population.un.org/household

9 Verity R, Okell LC, Dorigatti I, Winskill P, Whittaker C, Imai N, et al. Estimates of the severity of coronavirus disease 2019: a model-based analysis. Lancet Infect Dis. 2020; DOI:10.1016/S1473-3099(20)30243-7

